# SARS-CoV-2 Booster Effect and Waning Immunity in Hemodialysis Patients

**DOI:** 10.1101/2022.05.22.22275183

**Authors:** Eibhlin Goggins, Binu Sharma, Jennie Z. Ma, Jitendra Gautam, Brendan Bowman

## Abstract

**Background:** Dialysis patients are extremely vulnerable to SARS-CoV-2 infection. We recently reported the results of a prospective cohort study measuring serial monthly semi quantitative IgG antibody levels to the SARS-CoV-2 spike protein receptor binding domain in fully vaccinated in-center hemodialysis patients after receiving the BNT162b2 (Pfizer-BioNTech) mRNA vaccination.

**Methods:** Prospective cohort study measuring the serologic response of hemodialysis patients to a booster dose of BNT162b2 vaccine at an average of 2, 6 and 11 weeks post vaccination.

**Results:** Of 35 hemodialysis patients in the original cohort, 27 (77.1%) received a third dose of BNT162b2. Antibody level significantly increased from pre-booster to 2 weeks post-booster (median (25^th^, 75^th^ percentile) from 59.94 (29.69, 177.8) to 6216 (3806, 11730)), an average increase of 112 fold. Antibody levels dropped to a median of 2654 BAU/mL (1650, 8340) 6 weeks post-booster and to a median of 1444 BAU/mL (1102, 2020) between weeks 6 and 11 post-booster. Antibody levels at 11 weeks remained an average of 40 fold higher than pre-booster levels. Overall, antibody levels declined 47% month to month post-booster. Nine (33%) patients had negative or borderline detectable antibody levels pre-booster and 8 of 9 developed positive (>35.2 BAU/mL) antibody levels post-booster. Those with prior infection had a lower proportional increase in antibody level (51 fold) compared with the median change in COVID naïve patients (144 fold) from pre-booster to 2 weeks post-booster.

**Conclusions:** Our data demonstrates that hemodialysis patients obtain a robust humoral response from a third dose of the BNT162b2 vaccine although antibody levels wane over time.

---

Patients with End Stage Kidney Disease (ESKD) on dialysis suffer high morbidity and mortality from SARS-CoV-2. Despite successful vaccination campaigns by dialysis providers, the standard two-dose vaccination series with Pfizer BioNTech (BNT162b2) mRNA SARS-CoV-2 is insufficient to protect patients from infection due to the Omicron variant.^1^ Current guidelines recommend boosters of SARS-CoV-2 mRNA-based vaccines.^2^ However, data regarding humoral response post-booster is limited in dialysis patients. We previously reported long-term humoral responses to two doses of BNT162b2 in a cohort of hemodialysis patients.^3^ Six months after full vaccination, 40% of patients’ anti-spike protein IgG levels were either undetectable or borderline. Here, we report responses to a first booster of the BNT162b2 vaccine.

We performed a prospective cohort study measuring serial semi quantitative IgG antibodies to the SARS-CoV-2 spike protein S1 receptor binding domain. We evaluated the response at a mean of 2, 6 and 11 weeks post-booster. The Anti-SARS-CoV-2 QuantiVac ELISA (IgG) from Euroimmun (EUROIMMUN US, Inc.) was used in all assessments. Final results were reported in WHO-recommended binding antibody units (BAU/mL) per manufacturer’s instructions.^4^ Final results were considered negative for <25.6 BAU/mL, borderline for 25.6 to <35.2 BAU/mL and positive for ≥35.2 BAU/mL. Clinical data was obtained as previously described.^3^

Of 35 hemodialysis patients in the original cohort, 27 (77.1%) received a third dose of BNT162b2 and 20/27 (74%) had complete data (4 time point measurements): pre-booster (mean of 6 weeks pre-booster) and 2, 6 and 11 weeks post-booster. Differences in antibody levels over time were compared non-parametrically using the Friedman test, then tested pairwisely with Bonferroni correction at alpha of < 0.05. A linear mixed model was used to estimate decline in slope after vaccination. Antibody levels were significantly different over time (χ^2^(3)= 56.6, p<.0001). All pairwise tests were statistically significant. Antibody level significantly increased from pre-booster to 2 weeks post-booster (median (25^th^, 75^th^ percentile) from 59.94 (29.69, 177.8) to 6216 (3806, 11730)), corresponding to an average increase of 112 fold (Figure 1). Antibody levels dropped to a median of 2654 BAU/mL (1650, 8340) 6 weeks post-booster, a 36.3% average decrease. From weeks 6 to 11, there was a 52.4% drop to a median of 1444 BAU/mL (1102, 2020). Antibody levels at 11 weeks remained an average of 40 fold higher than pre-booster levels. Overall, antibody levels declined 47% month to month post-booster.

**Figure 1:**
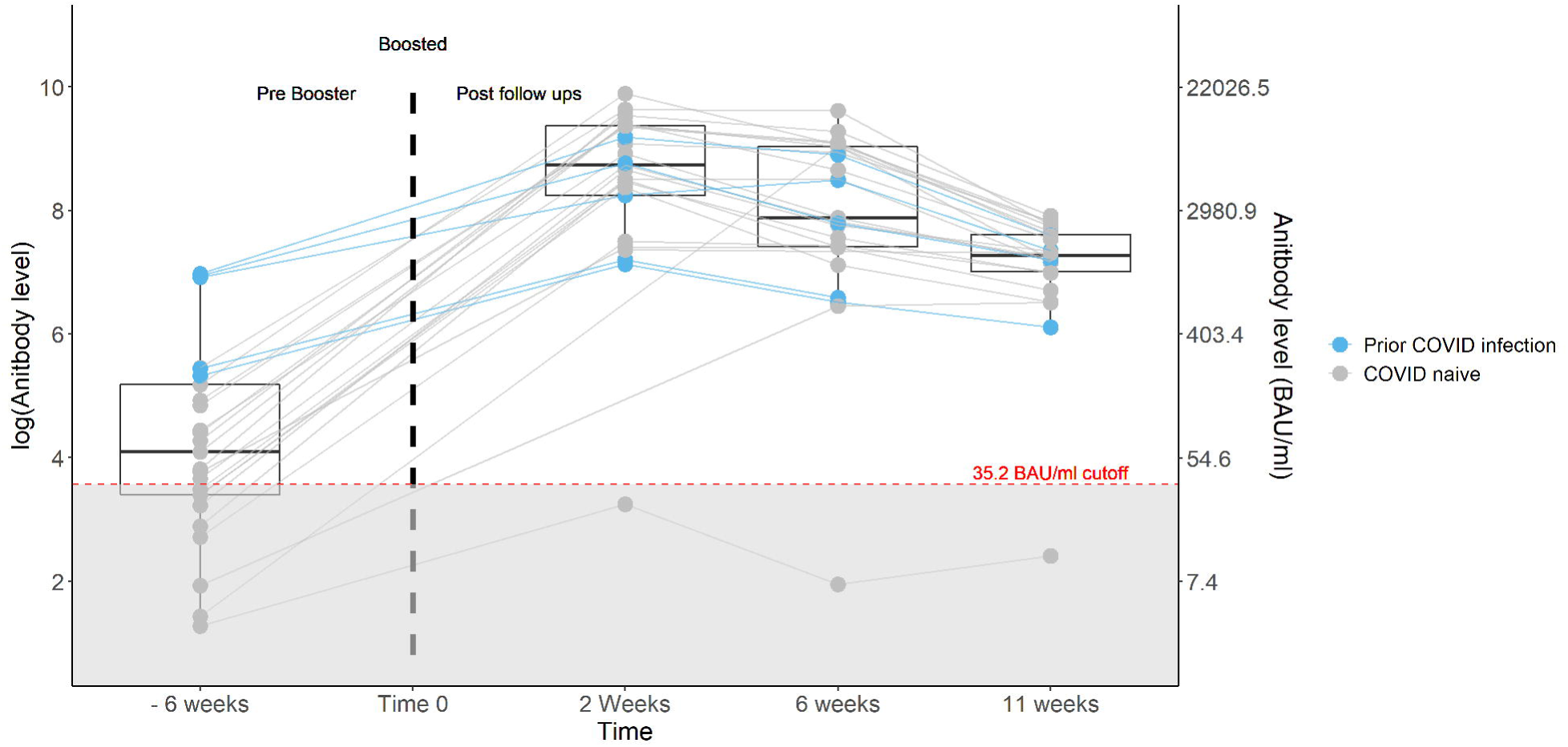
Antibody levels before and at an average of 2, 6 and 11 weeks after a SARS-COV-2 BNT162b2 booster vaccine in 27 patients. Each dot represents a subject and each line represents a single patient trend. The median level is represented by the horizontal line in the box at each time point. Dots under the red dashed line were below the borderline/negative cutoff for antibody level protection.

Nine (33%) patients had negative or borderline detectable antibody levels pre-booster and 8 of 9 developed positive (>35.2 BAU/mL) antibody levels post-booster. Notably, the one patient not achieving positive antibody levels was immune suppressed. Six patients had prior SARS-CoV-2 infection as identified by a Bio-Rad Platelia SARS-CoV-2 Total Ab assay assessing anti-nucleocapsid IgG antibodies (Bio-Rad Laboratories, Inc.). From pre-booster to 2 weeks post-booster, those with prior infection had a lower proportional increase in antibody level (51 fold) compared with the median change in COVID naïve patients (144 fold).

In earlier results, 40% of the cohort transitioned from positive to negative antibody status at 6 months post two-dose vaccination. Encouragingly, a third dose appears to restore antibodies to high levels, though these waned quickly in ensuing weeks. Similar trends of antibody decline are seen in healthy individuals, although dialysis patients may differ from the general population with reduced peak levels and lower seroconversion rates.^5^ Long term durability remains unclear and protective levels against infection are unknown. Goldblatt et al. reported the mean protective threshold against WT SARS-CoV-2 virus was 154 BAU/ml (95 % CI 42–559) but higher levels are presumed required against current variants.^6^

Interestingly, previously infected patients saw a blunted rise in antibodies to an initial booster shot, though these patients started from a higher baseline. Thus, overall, they attained similar peak levels. While our sample size precludes further analysis of this finding, the interaction of natural immunity with booster vaccination response requires further study.

During the recent Omicron wave, boosters were found to be protective from hospitalization and severe illness in the general population; however, this effect was time dependent and declined significantly at four months post-booster.^7^ A similar pattern is likely in patients on dialysis, but few studies have been conducted in this population. In one study, 93% of dialysis patients who received a third dose of BNT162b2 vaccine achieved antibody levels associated with protection, compared with only 35% pre-booster.^8^ Recently, the Centers for Disease Control recommended a fourth dose of mRNA vaccine for select populations.^2^ The utility of such a strategy in dialysis patients remains unclear but the humoral antibody waning seen in dialysis populations may support additional boosters.

Our study has limitations: small sample size, brief follow up time and focus on humoral immunity. In conclusion, our data illustrates that, although humoral immunity wanes, patients on hemodialysis demonstrate strong antibody responses to a third dose of the BNT162b2 vaccine.

## Supporting information

COI_Disclosure_BB

COI_Disclosure_JG

COI_Disclosure_EG

COI_Disclosure_JM

COI_Disclosure_BS

StrobeChecklist

## Data Availability

Data is not freely available externally. Please contact author if interested in de-identified data set for research purposes

## Ethics approval and consent to participate

This research proposal was reviewed and approved by the University of Virginia Institutional Review Board for Health Sciences Research (tracking number: HSR 210095).

## Availability of data and materials

The dataset used for this analysis is not publicly available. The data utilized was obtained from the Electronic Health Record and from the dialysis-specific electronic medical record system, which is restricted to use by only authorized employees.

## Acknowledgements

The authors gratefully acknowledge the following individuals for their contributions to this study. First, Elizabeth Kirkland for providing IRB and study support as lead clinical research coordinator. Danielle Wentworth MSN, FNP for assistance consenting and obtaining samples. The UVA dialysis center managers and staff who assisted in obtaining monthly samples. Drs. Mark Okusa and Julia Scialla for providing study design feedback and support. And most importantly, our patients who agreed to join our study.

